# A systematic review about skin lesions in children affected by Coronavirus Disease 2019 (excluding multisystem inflammatory syndrome in children): study protocol

**DOI:** 10.1101/2022.03.04.22271888

**Authors:** Arianna Dondi, Giacomo Sperti, Davide Gori, Marco Montalti, Lorenza Parini, Federica Guaraldi, Marcello Lanari, Iria Neri

**Affiliations:** Pediatric Emergency Unit, Scientific Institute for Research and Healthcare (IRCCS), Azienda Ospedaliero-Universitaria di Bologna, Bologna, Italy; Specialty School of Pediatrics, Alma Mater Studiorum, University of Bologna, Bologna, Italy; Department of Biomedical and Neuromotor Sciences, University of Bologna, 40126 Bologna, Italy; School of Hygiene and Preventive Medicine, Department of Biomedical and Neuromotor Sciences, Public Health and Medical Statistics, University of Bologna, Via San Giacomo 12, 40126 Bologna, Italy; IRCCS Istituto delle Scienze Neurologiche di Bologna, 40139, Bologna, Italy; Dermatology Unit, Scientific Institute for Research and Healthcare (IRCCS), Azienda Ospedaliero-Universitaria di Bologna, Department of Experimental, Diagnostic and Specialty Medicine, University of Bologna, Bologna, Italy

**Author notes:** <u>Corresponding author</u>: Davide Gori, Department of Biomedical and Neuromotor Sciences, University of Bologna, 40126 Bologna, Italy.

## Abstract

COVID-19 disease can give a range of skin manifestations, some of which specific to children and young patients. Given the different nature of the lesions in specific stages of the disease and the fact that they may be the only or predominant symptom of the disease, it is of great importance for the pediatrician and dermatologist to be familiar with COVID-19 skin lesions. The aim of this systematic review, conducted according to the PRISMA statement, is to investigate COVID-19-associated cutaneous involvement in children in terms of type of skin lesions, frequency, and time of onset, with the exclusion of those associated with multisystem inflammatory syndrome in children. A comprehensive literature search will be performed through PubMed between December 2019 and December 2021. The quality of each study will be assessed according to the STROBE tool for observational studies. This systematic review will provide evidence about the dermatological manifestations of COVID-19 disease in children published up to the end of year 2021. Recognizing skin symptoms as potential manifestations of COVID-19 in children is important for a non-delayed diagnosis, with prompt activation of the adequate tracing, isolation, and hygienic measures, and for timely treatment of unlikely but possible serious complications.

## Introduction

The first case of infection by the Severe Acute Respiratory Syndrome Coronavirus 2 (SARS-CoV-2), causing an illness called Coronavirus disease 2019 (COVID-19), was recognized in China in December 2019.(1) Since then, the virus rapidly spread to cause a pandemic. The clinical manifestations of COVID-19 are diverse, extending from mild symptoms to critical lethal respiratory failure and septic shock. In the pediatric age, the disease is usually milder than in adults, even if rare but potentially severe consequences such as the Multisystem Inflammatory Syndrome in Children, a rare hyperinflammatory condition that can occur some weeks after the infection(2) or the long-COVID-19 syndrome(3) may arise.

A constellation of cutaneous manifestations have been observed in COVID-19. Some of them, such as urticaria, maculopapular or vesicular rashes, can occur at any age, but certain manifestations such as chilblains, erythema multiforme are more frequently seen in children and young patients.(4) Moreover, different skin manifestations have been described at different stages of the disease, and some of them might be the only or more prominent presentation of COVID-19.

It is therefore mandatory for the pediatrician and the dermatologist to be familiar with the COVID-19-associated skin lesions for a prompt diagnosis and a correct management of the spread of the infection. The aim of this systematic review is to provide evidence about COVID-19-associated cutaneous involvement in children in terms of type of skin lesions, frequency, and time of onset. The skin lesions commonly associated with MIS-C were excluded from this work because they can be considered as part of the spectrum of this severe complication of COVID-19 and may deserve to be dealt with separately.

## Methods/Literature Search

We will conduct a systematic review, following the Preferred Reporting Items for Systematic Reviews (PRISMA) approach(5). Literature search will be performed on PubMed by using the following string for a thorough search: “(((SARS-CoV-2) OR (COVID19) OR (COVID-19) OR (ncov*) OR (coronavirus)) AND ((Child) OR (children) OR (pediatric) OR (paediatric) OR (infant) OR (adolescent)) AND ((“2019/12/31”[Date - Entry] : “2021/12/12”[Date - Entry]))”.

We will include full-text accessible Italian and English-language articles; population: children and young people aged <18 years; study design: observational prospective or retrospective studies, case series, case reports; outcome: any skin lesions in patients affected by COVID-19.

We will exclude case reports in which COVID-19 is not confirmed by the diagnostic procedures (molecular or antigenic swab, serology, or RT-PCR on bioptic specimen); any study in which the dermatologic presentation is vague (e.g. “cutaneous lesions”, “any dermatological lesion”).

Studies involving both pediatric and adult patients will be included if children and adolescents’ characteristics can be clearly discernible or if this age category represents a meaningful part of a group/cohort and the mean or median age is <18 yrs.

### Data Extraction

Data will be extracted by six independent reviewers. Two authors will independently retrieve each title from the searches. On the basis of the title and abstract, eligibility for the article will be determined, and the full text of the selected papers will provide information for the final decision of inclusion or exclusion. A manual search will also be performed by reviewing bibliography of pertaining articles to identify additional studies.

The pertinent selected literature will hence be tabulated with regards to study design, study setting, number of patients, sex, age, phenotypic traits, COVID-19 diagnosis (nasopharyngeal molecular or antigenic swab, serology, RT-PCR on skin lesions), type of skin lesions, time of onset of skin lesions with respect to COVID-19, diagnostics on skin lesion (histology or electronic microscopy) and its timing, other symptoms, other skin lesions, comorbidities or risk factors, treatment for COVID-19, treatment for skin lesion, other treatments, follow-up length, outcome.

### Quality Assessment

Two authors will independently and blindly assess the quality of the included studies using the “Strengthening the Reporting of Observational Studies in Epidemiology” (STROBE) tool for observational studies(6). Any disagreement between the two researchers will be resolved through discussion. If discussion will not be sufficient, a third blind reviewer will appeal as tie breaker. The STROBE statement is a 22 items tool specifically designed to evaluate observational studies quality. 18 items are the same in the three different checklist and five questions (6-12-14-15) are differently formulated for each study design: 1) Cohort study, 2) Case report study, 3) Cross sectional study. STROBE does not provide ways to clearly define a score allowing to rate the quality of the study. As a general rule, the higher is the score, the higher is the quality of the study. We will use the cut offs for three levels of score: 0-14 as poor quality, 15-25 as intermediate quality and 26-33 as good quality of the study(7).

## Results and discussion

### The results of the literature search will then be analyzed and discussed

Our systematic review will provide evidence about all the dermatological manifestations of COVID-19 in children published up to the end of year 2021. After two years since the beginning of the pandemic, it is well-known that COVID-19 can have variable and multiform clinical presentations, and that skin is often affected. Children are usually less severely affected than adults, however they can spread the disease to at-risk groups; moreover, some of them can have serious complications as well. Recognizing skin symptoms as potential manifestations of COVID-19 in children is important for a non-delayed diagnosis, with prompt activation of the adequate tracing, isolation, and hygienic measures, and for timely treatment of unlikely but possible serious complications.

## Data Availability

All data produced in the present work are contained in the manuscript.

